# Composite proteomic and metabolomic plasma biomarkers for detection of colorectal, lung and ovarian cancers

**DOI:** 10.1101/2025.05.19.25326282

**Authors:** Jim Åkerrén Ögren, Joakim Ekström, Natallia Rameika, Emma Torell, Chatarina Larsson, Gunilla Enblad, Ivaylo Stoimenov, Patrick Micke, Ulf Gyllensten, Mats Hellström, Bengt Glimelius, Karin Stålberg, Tobias Sjöblom

## Abstract

Sensitive and specific blood biomarkers for detection of cancer are highly warranted. To discover such biomarkers, we measured plasma levels of 165 proteins and 244 metabolites in 818 patients with colorectal, lung or ovarian carcinoma at diagnosis, 119 patients with non-malignant conditions of the corresponding organs, and 1,129 healthy individuals. We performed exhaustive search over all cut-off values of the ROC statistic and identified composite biomarkers with diagnostic performance significantly superior to benchmark FDA approved blood tests in clinical use for detection of cancer. We found biomarkers composed of 2-4 proteins separating cases of each tumor type from healthy controls with ROC AUC in the range 0.89 to 0.98. These biomarkers also separated cases of each tumor type from the other two (ROC AUC 0.82-0.88). For lung and ovarian cancers, we identified biomarkers distinguishing cases with intermediate and high from those with low tumor stages. These biomarkers for cancer detection and stage can find use in early detection, staging and differential diagnosis of common tumor types.

## Introduction

Colorectal cancer (CRC), lung cancer (LuCa) and ovarian cancer (OvCa) are among the cancers with the highest global incidence, with LuCa and CRC placing 1st and 3^rd^ for all cancers, and OvCa being the 8^th^ most diagnosed cancer in women^1^. Lung cancer and CRC are also the two leading causes of cancer-related death, accounting for 18.7% and 9.3% of all deaths from cancer, respectively, and in females OvCa places 8^th^ with 4.8% of deaths^1^. In most common cancer types, stage at diagnosis is an important predictor of survival^2, 3, 4^. Diagnosing the tumor at an early stage is generally perceived to increase access to curative surgery for the primary tumor, but in e.g. the US, nearly one-quarter of CRC, half of LuCa and up to 70% of OvCa cases are currently diagnosed at a late stage with distant metastases present^3^. Screening programs may contribute to early detection and diagnosis at earlier stage^5^ by pinpointing tumors before development of symptoms, and are shown to reduce the mortality in CRC^6^ and LuCa^7, 8^. In combination with ultrasound, CA125 as a single biomarker was recently found insufficient to support OvCa screening in an asymptomatic population as it did not significantly lower the mortality although more cancers were diagnosed at lower stages^9^. The symptoms associated with CRC, LuCa, OvCa and several other malignancies are often non-specific and partially overlapping at earlier stages, including fatigue, weight loss, abdominal pain, constipation or diarrhea. There is therefore a need for sensitive and specific blood tests that detect these tumor types at early stages and also discriminate them from each other.

Circulating biomarkers offer potential for non-invasive diagnostic and screening tools in cancer. Blood testing is safe and associated with high patient adherence compared to other methods such as radiology, endoscopy or stool-based tests. Current large-scale molecular analysis techniques enable parallel measurement of thousands of analytes in small volumes of blood, including e.g. levels, variants and epigenetic or post-translational modifications of proteins, nucleic acids and metabolites. Recent advances in pan-cancer blood biomarker tests include combined detection of proteins with mutational analysis of ctDNA^10^ and analysis of cfDNA methylation with/without fragmentation^11, 12, 13, 14^. These efforts have led to ongoing prospective clinical trials that compare cancer incidence rates in conventional screening methods with blood-based screening tests, exemplified by the NHS-Galleri Trial^15^, PATHFINDER 2 (NCT05155605), STRIVE (NCT03085888), and SUMMIT (NCT03934866) studies. While the specificity of methylation-based tests is high, sensitivity is lower particularly in early tumor stages for several common tumor types, potentially because of low levels of released ctDNA.

To ensure efficient translation, biomarker discovery should ideally align with the efficacy requirements from regulatory authorities that govern their use as diagnostic tests, such as the US Food and Drug Administration (FDA). In practice, for a novel biomarker to gain regulatory approval as a medical device for clinical use, non-inferior ROC AUC to an existing approved test for the same purpose must be demonstrated. Diagnostic blood biomarkers of cancer currently in clinical use generally suffer from low sensitivity and/or specificity, which limits their utility. In CRC, blood tests could improve participation rates in screening programs that are currently based on fecal samples^16, 17^, however the current best FDA approved blood test to discover CRC, Epi proColon (ROC AUC 0.82)^18, 19^, does not reach the performance of either the fecal occult blood test (FIT) alone (ROC AUC 0.88)^20, 21^, or the multi-target stool DNA test Cologuard (ROC AUC 0.93)^22, 23^. Notably, across all other cancer types, no blood test with performance on par with Epi proColon for CRC is currently FDA approved for early detection. Taken together, there is a need for robust blood biomarker tests for cancer with diagnostic performance exceeding the state-of-the-art.

Here, we analyze plasma proteome and metabolome data using a ROC-based biomarker discovery approach with benchmarking to regulatory approved early detection tests for cancers to find composite diagnostic plasma biomarkers that have suitable properties for further development.

## Results

### Plasma proteomic and metabolomic analyses of cases and controls

To discover diagnostic biomarkers, we measured the levels of 165 proteins and 244 metabolite parameters in plasma samples drawn at diagnosis before any treatment from 330 CRC, 304 LuCa, and 184 OvCa (including 24 borderline tumor) patients with tumor stages I-IV from the prospective population-based cancer biobank U-CAN^24^ (Figure 1A, Supplementary Figure 1). We also analyzed blood plasma from 119 (50 for CRC, 16 for LuCa and 53 for OvCa) patients with non-malignant diagnosis from the same biobank, here termed Non-cancer controls, to enable selection of biomarkers that discriminate cancer from pre-malignant or non-malignant diseases of the same organs (Figure 1B, Supplementary Figure 2). As healthy population controls, we analyzed plasma samples from 1,129 individuals of the EpiHealth cohort^25^ that were in the same age span as the cancer cases and living in the same area (Figure 1B, Supplementary Figure 1). To ensure a relevant age interval for cancer screening, the cases and controls were of age 45-75 in CRC and LuCa and 40-80 in OvCa. The age and sex distributions of cases and population controls were similar (Table 1, Figure 1B). Comparing the study cohort to the incident population as recorded in the national quality registries for the respective cancers, we found that for CRC, the distributions of patient sex, tumor location, T and N stage was similar to the incident population (Supplementary Figure 3). In LuCa, the proportions of smokers, sex and histological subtype were similar, whereas stage I was overrepresented and stage IV underrepresented compared to the incident population (Supplementary Figure 4). For OvCa, the relative proportion of sub-diagnoses differed, and stage I tumors were underrepresented compared to the population, but there was a similar ratio of stages I-III to IV/X0 (Supplementary Figure 5). Together, the population-based study cohorts are representative of the incident cancer cases and the background population.

**Figure 1.**
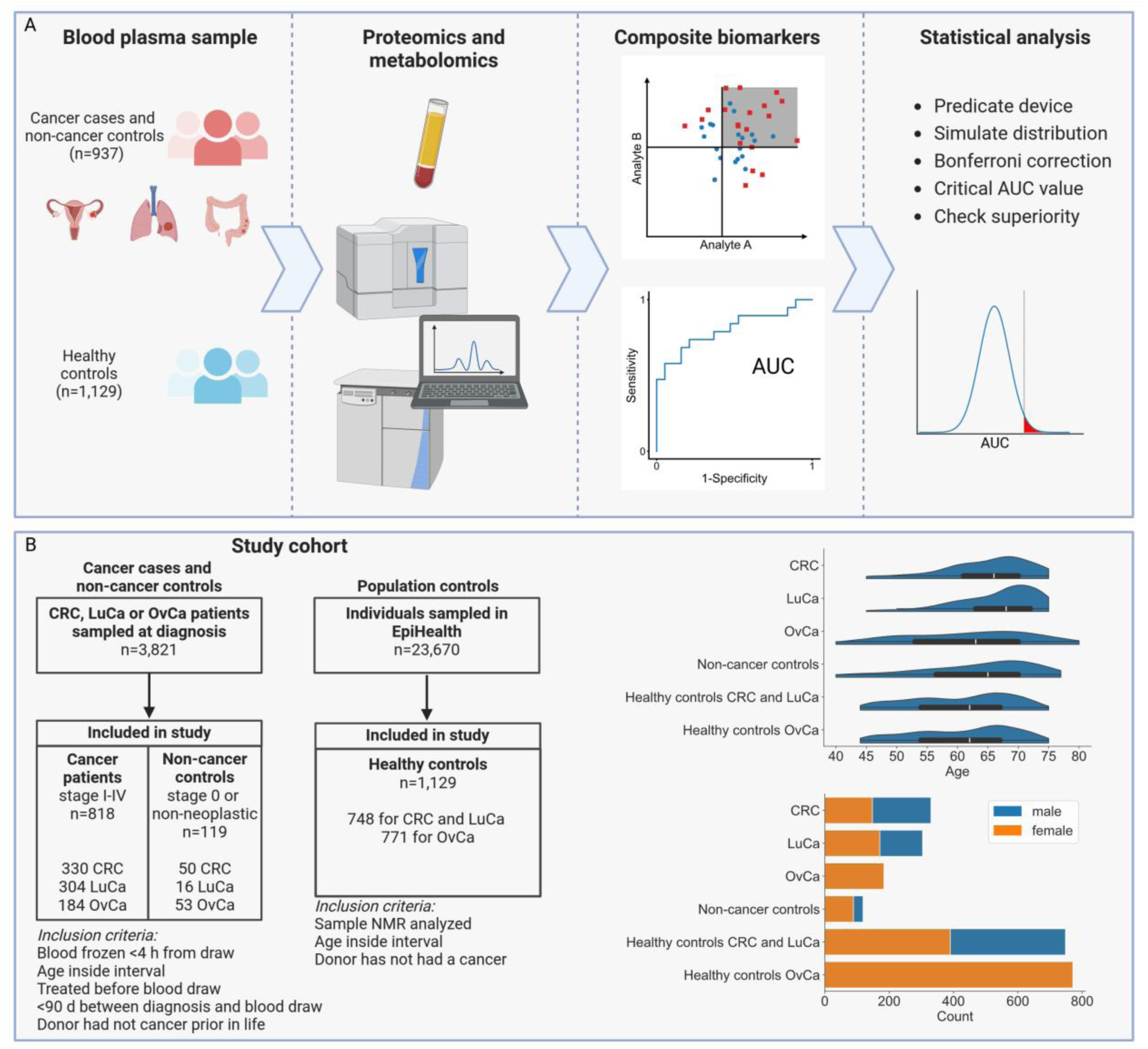
Discovery of tumor-type specific and pan-cancer biomarkers in blood plasma. A. Plasma samples from patients with cancer or non-malignant conditions at diagnosis and healthy population controls were analyzed by affinity proteomics and NMR metabolomics. The ROC was computed for individual analytes and for composite biomarkers of 1-4 analytes within and across the study cohorts. The ROC AUC distributions for biomarkers with performance similar to a predicate device already in clinical use were used for statistical testing of putative biomarkers. B. Selection of cases and controls and characteristics of the selected study populations. From 3,821 cases being investigated for colorectal, lung and ovarian cancer, sampled for blood at time of diagnosis in the population-based prospective study U-CAN, 818 with confirmed cancer and 119 with pre-malignant tumors or other disease of the respective organs fulfilled the inclusion criteria. From 23,670 healthy individuals in the population-based cohort EpiHealth, 748, and 771 controls were selected for the CRC and LuCa, and the OvCa cohorts, respectively.

**Table 1.**
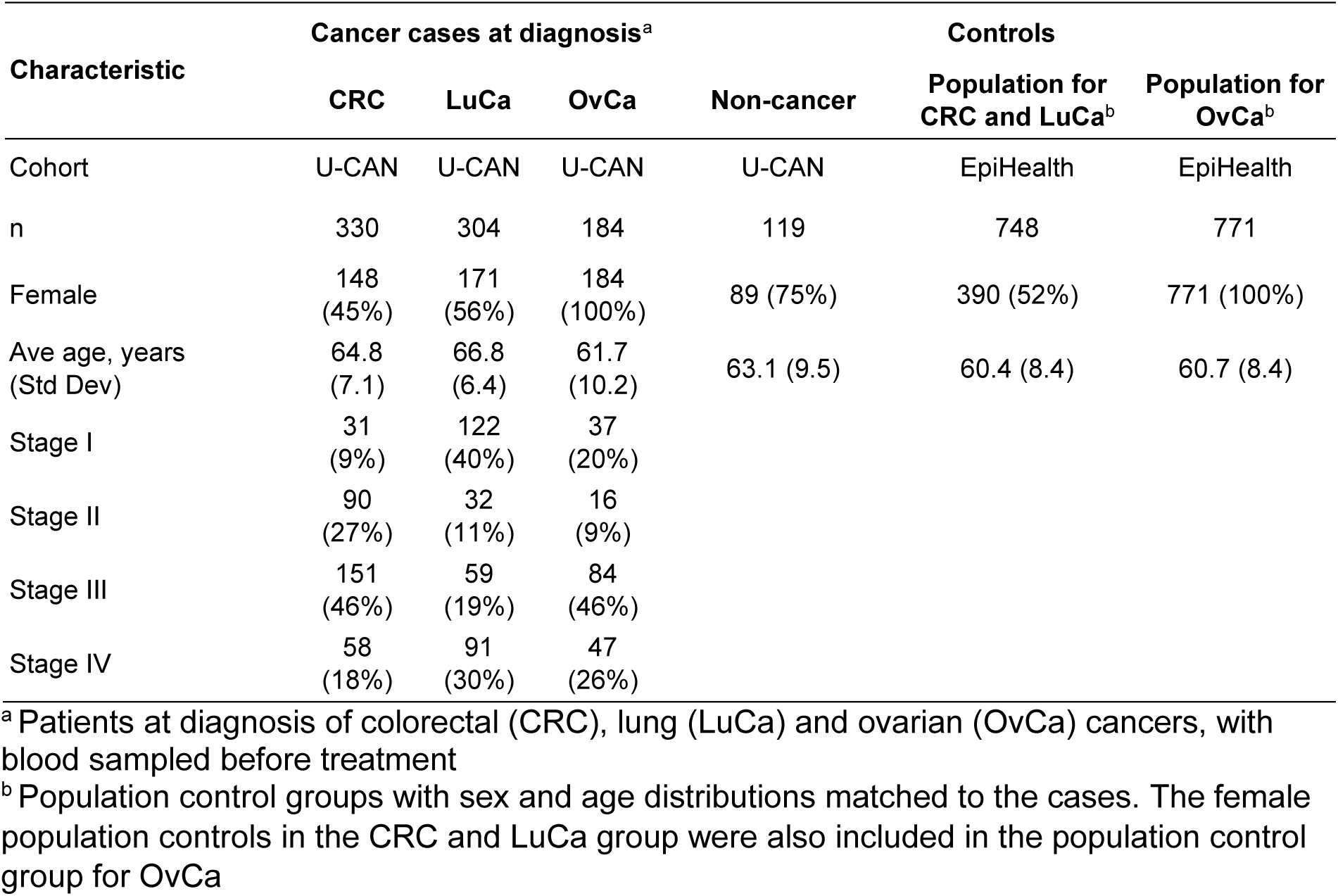
Characteristics of cancer cases and controls.

### Protein and metabolite levels altered in cancer patients versus non-cancer controls

Since the cancer cases and healthy population controls were from different biobank cohorts, we expected that there could be pre-analytical effects that could affect biomarker identification. Indeed, we observed that the vast majority of NMR analytes discriminated between non-cancer controls and healthy controls, whereas only a subset of proteins did. In an attempt to minimize the impact of pre-analytical variation, we first analyzed the cancer cases versus the non-cancer controls from the same cohort, before proceeding to comparing cancer cases to the population controls. First, we computed the ROC AUC for each measured protein as single biomarker in discriminating cancer cases from the 119 non-cancer controls drawn from the U-CAN cohort, and tested for statistical significance (Figure 2A). From the 165 proteins measured, 22 (CRC), 31 (LuCa), 12 (OvCa) and 14 (pan-cancer) were up- or downregulated in plasma from the cancer patients at diagnosis (p < 0.05, 2-sided Mann-Whitney *U* test under Bonferroni correction; Supplementary Tables 1-5). Several established biomarkers for each tumor type were corroborated, including (1) ferritin down-regulation in CRC (AUC 0.66)^26, 27^, (2) carcinoembryonic antigen (CEA/CEACAM5) up-regulation with a ROC AUC of 0.68 and 0.69 for CRC and LuCa, respectively, congruent with AUC 0.67-0.80 for CRC^10, 28, 29, 30, 31, 32, 33, 34, 35^ and 0.62-0.82 for LuCa^10, 36, 37^, (3) HMGB1 up-regulation in LuCa (AUC 0.76)^38^, (4) SPP1/OPN up-regulation in all three tumor types^10^, and (5) Mucin 16 (CA125) with ROC AUC 0.91 for OvCa, in the reported range 0.77-0.97^10, 39, 40, 41, 42, 43, 44, 45, 46^ (Figure 2B). Further, novel potential biomarkers were discovered, including PECAM1/CD31 up-regulation in CRC (AUC 0.69) and LuCa (AUC 0.68). Investigating the overlap between diagnoses, we found that seven significantly regulated proteins connected CRC and LuCa, 3 connected LuCa and OvCa, but none connected CRC and OvCa (Figure 2C).

**Figure 2.**
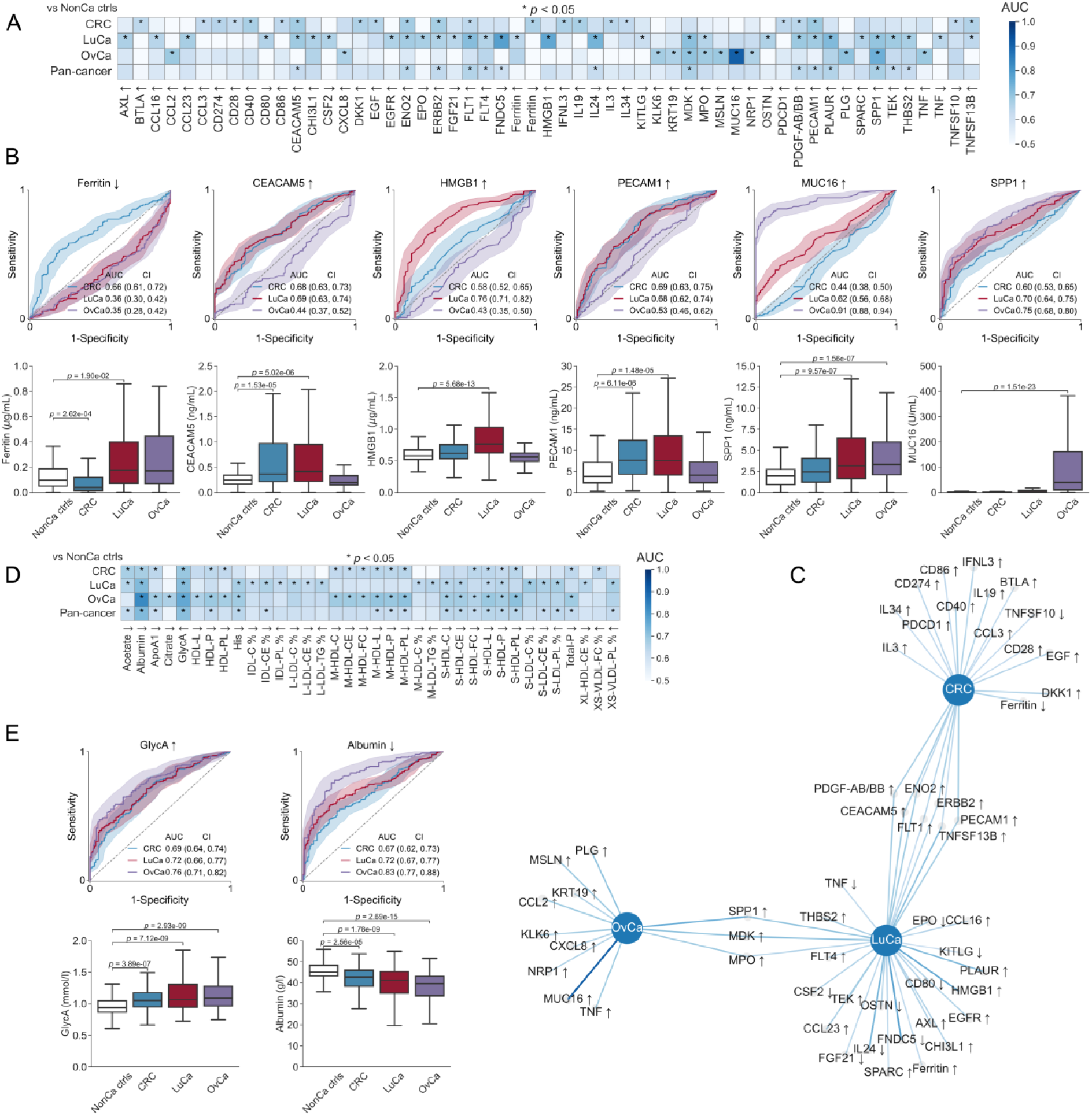
Pan-cancer and tumor-type-specific protein and metabolite biomarkers. For each cancer type alone and together (“pan-cancer”), ROC AUC values were computed for all 165 proteins and 244 metabolites comparing cancer cases against 119 non-cancer controls within the U-CAN cohort. A. ROC AUC values for proteins significantly up- or down-regulated for at least one cancer type. * denotes significant analytes at the 95% level (p < 0.05). B. ROC and Box plots for down-regulated Ferritin in CRC, up-regulated CEACAM5 (CEA) in CRC and LuCa, up-regulated HMBG1 in LuCa, up-regulated PECAM1 in both CRC and LuCa, up-regulated SPP1 (Osteopontin) in LuCa and OvCa, and up-regulated MUC16 (CA-125) in OvCa. All *p* values adjusted using two-sided Mann-Whitney *U* test under Bonferroni correction for multiple hypothesis testing. Confidence bands in ROC plots contain 95% of ROC curves from 1,000 bootstrap iterations. C. Graph visualization of proteins regulated at 95% significance. D. ROC AUC values for metabolites significantly up- or down-regulated for at least one cancer type. E. ROC and Box plots for pan-cancer markers up-regulated GlycA and down-regulated albumin.

Next, we computed the ROC AUC for each NMR determined analyte in discriminating cancer cases from non-cancer controls from the same cohort and tested for statistical significance. From the 244 determined metabolites, 18 (CRC), 21 (LuCa), 21 (OvCa) and 20 (pan-cancer) were significantly up- or downregulated in cancers (p < 0.05, 2-sided Mann-Whitney *U* test under Bonferroni correction; Figure 2D, Supplementary Tables 1-5). Several of these were supported by previous studies. Reduced albumin^47^, increased levels of the systemic inflammation marker glycoprotein acetyls (GlycA)^48, 49^ along with reduced S-HDL-L and S-HDL-P were potential biomarkers of all three tumor types (Figure 2E). The CRC and OvCa groups had reduced lipoprotein particles and ApoA1^50^ as well as reduced concentration and altered composition of HDL particles, along with reduced total concentrations of lipoprotein particles (Total-P), phospholipids (Total PL), and free cholesterol (Total FC). The LuCa cases had altered LDL composition with reduced cholesteryl esters and cholesterol fraction and increased phospholipids fraction.

### Composite biomarkers for separation of cancer patients at diagnosis from healthy population controls

We next proceeded to identify composite biomarkers that could discriminate cancer cases from healthy population controls. To remove false positive biomarkers arising from pre-analytical differences between the cohorts, we included only the proteins and metabolites that by themselves could discriminate the cancer cases from the non-cancer controls (Figure 2A and D). We here required these individual analytes to also have significant difference versus the population controls. Finally, to further ensure that we filtered analytes with large pre-analytical or cohort effects, we retained only those where ROC AUC for cancer vs non-cancer controls was greater than ROC AUC for non-cancer controls vs healthy population controls. This resulted in 10 analytes (10 proteins, 0 metabolites) for CRC, 22 (21 proteins and 1 metabolite) for LuCa, and 13 (12 proteins and 1 metabolite) for OvCa identified as suitable for being part of composite biomarkers for the respective cancer type (Table 2, Supplementary Figure 6, Supplementary Tables 6-9).

**Table 2.**
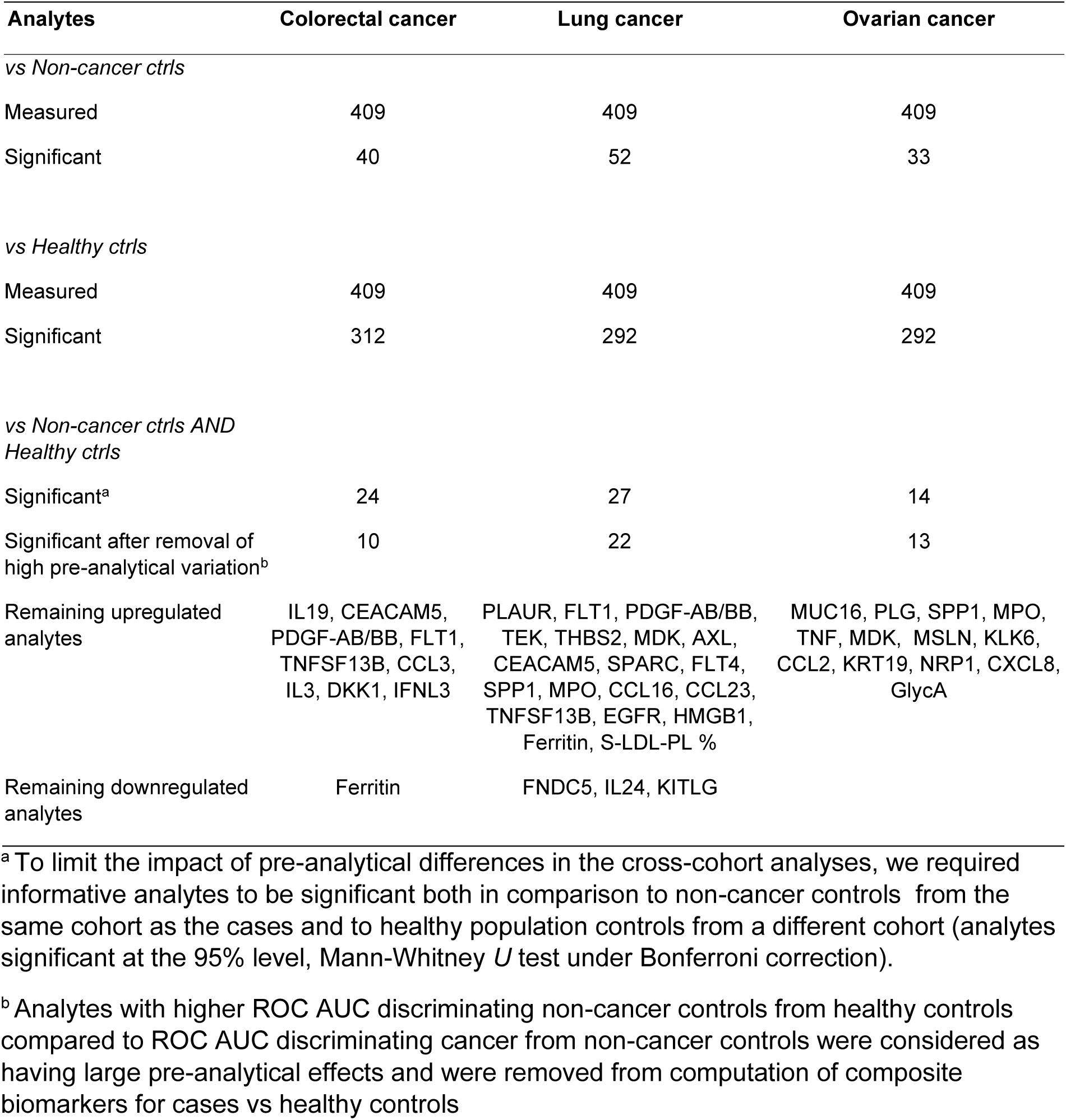
Protein and metabolite analytes with statistically significantly altered levels in blood plasma from cancer cases compared to controls.

When forming composite biomarkers for each tumor type, we evaluated each analyte up- or downregulated and all analytes combined using logical AND or logical OR resulting in *2^k+1^* classification scenarios where *k* denotes the number of analytes combined (Supplementary Figure 7). For each classification scenario we performed exhaustive search on all possible combinations of cutoff values to obtain the optimum ROC performance, i.e. finding the best true positive rate for each false positive rate. It can be argued that only biomarkers that supersede existing regulatory authority approved and clinically used diagnostic tests are meaningful to discover. We therefore evaluated all combinations of 2-4 up- or downregulated analytes, computed the ROC AUC for different classification scenarios and performed statistical hypothesis testing for superiority versus biomarkers with performance on par with regulatory approved diagnostic tests. For benchmarking purposes, we used the CRC tests Epi proColon/ColoHealth (AUC 0.82), FIT (AUC 0.88) and Cologuard (AUC 0.93)^51^ throughout, arguing that performance of AUC 0.82 is a minimal performance target also for early detection of OvCa or LuCa. The FIT and Cologuard tests are feces-based, but they represent state-of-the-art tests for early detection of cancer with performance levels that new biomarkers will need to match or supersede for regulatory approval and adoption in clinical use.

For each tumor type we computed all possible composite biomarkers combining 2, 3 and 4 analytes and checked for superiority against these three predicate device tests. For CRC, there were only 2 composite biomarkers of 4 analytes that were significantly superior to a test on par with Epi proColon, while for LuCa, there were 3 composite biomarkers of 2 analytes, 60 of 3 analytes, and 783 of 4 analytes that met the benchmark. For OvCa there were many composite biomarkers significantly better than a test with performance on par with Cologuard (Table 3, Supplementary Tables 10-17). For the best biomarkers per tumor type, a combination of upregulated CEACAM5 (CEA), FLT1 and IL19, and downregulated Ferritin separated CRC from healthy population controls (AUC 0.89), and a combination of downregulated FNDC5 and upregulated MDK, PLAUR and CEACAM5 separated LuCa from healthy population controls (AUC 0.91) (Figure 3B). Both these biomarkers were statistically significantly superior to a biomarker on par with the CRC blood test Epi proColon but not to a biomarker on par with FIT. A combination of upregulated MUC16 (CA125) and PLG separated OvCa from healthy population controls (AUC 0.97) with performance significantly superior to a test with performance on par with Cologuard. Excluding CA125, which performed exceptionally well here as compared to the literature, a combination of upregulated PLG, KLK6, MDK and CCL2 separated OvCa from healthy population controls with AUC 0.92, which was significantly superior to a biomarker with performance on par with Epi proColon. This performance is comparable to that of the OvCa test OVA1, albeit OVA1 is not approved for screening. The best biomarkers for each tumor type could also separate cancer cases from the non-cancer controls with AUC 0.81-0.92 and discriminate the target tumor type from the two other tumor types with AUC 0.82-0.88 (Figure 3A). Closer analysis of the best biomarkers per tumor type revealed that all the individual analytes contributed to the performance of the respective composite biomarker (Figure 3B). Further, the composite biomarkers discriminated later stage tumors from healthy controls better than they discriminated earlier stages from healthy controls (Figure 3C), which was also the case for the individual analytes included in the combinations (Supplementary Figure 8). All of the top-performing composite biomarkers were determined from plasma protein levels only (Figure 3D), none included metabolites. Replacing upregulated MDK protein with upregulated metabolite S-LDL-PL % in the four-combination for LuCa resulted in slightly worse but comparable ROC AUC (Supplemental Table S13) but overall the benefit of adding a fourth analyte to the three-protein core of PLAUR, FNDC5 and CEACAM5 was small. Similarly, the metabolite analyte GlycA was included in significant OvCa biomarkers but only as marginal improvements to a strong core based on protein combinations. Taken together, we discovered two-protein biomarkers for OvCa and four-protein biomarkers for CRC and LuCa that were superior to relevant benchmark FDA approved blood and/or stool tests for early diagnosis of cancer.

**Figure 3.**
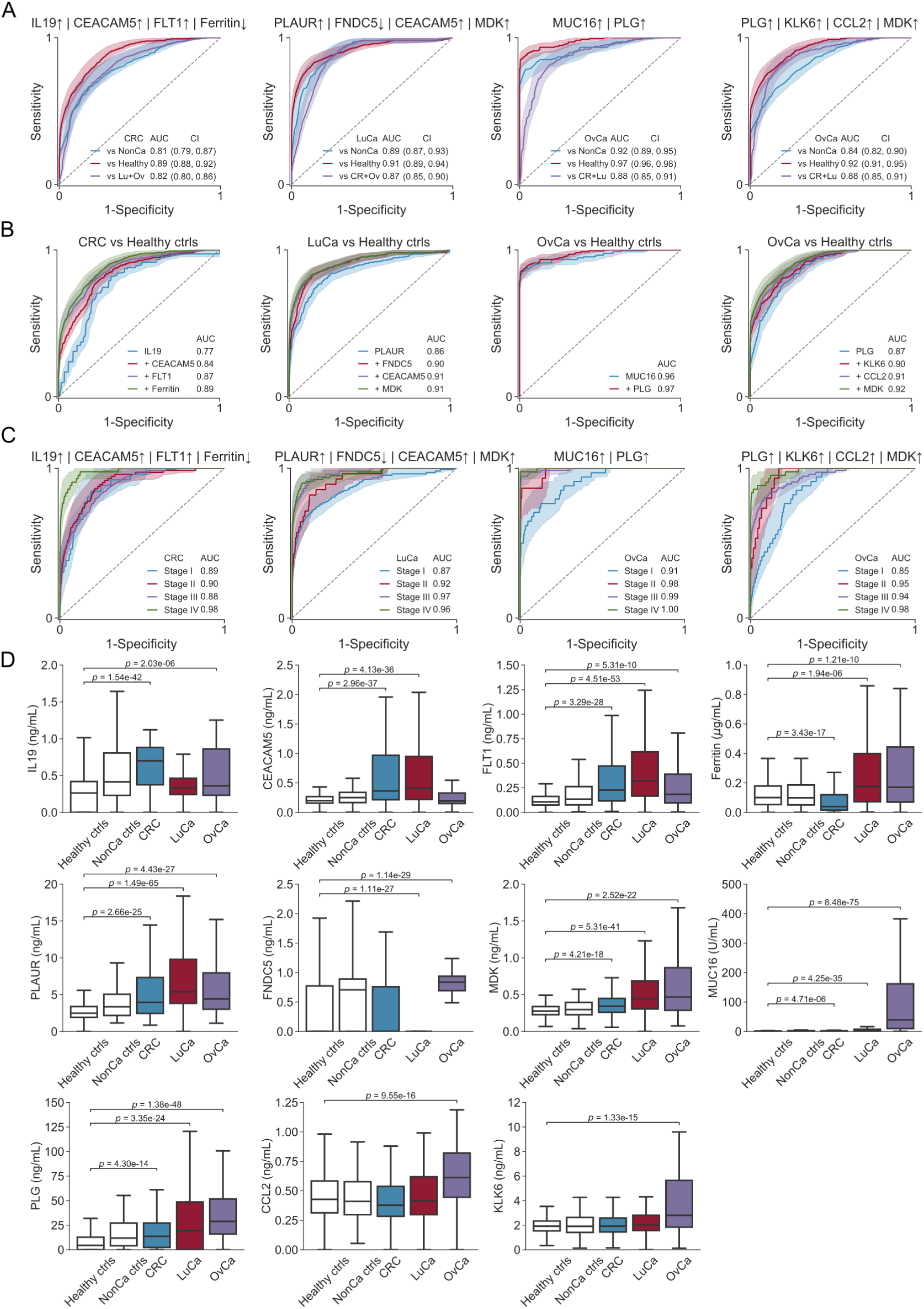
Composite biomarkers distinguish patients with either CRC, LuCa or OvCa from healthy individuals and from patients with the two other forms of cancer. The ROC was computed for all combinations of 1-4 of the selected analytes for each cancer type. For each combination evaluated, we optimized the ROC curve by exhaustive search on all possible cutoff values combined either by logical OR (|) or logical AND (&). The resulting composite biomarker was evaluated for superiority with respect to Epi proColon (AUC 0.82), FIT (AUC 0.88) and Cologuard (AUC 0.93) at the 99% significance level using Bonferroni correction for multiple hypothesis testing. A. ROC plots with AUC values for the top composite biomarkers for colorectal cancer (CRC), lung cancer (LuCa), ovarian cancer (OvCa), and OvCa when CA125/MUC16 was excluded from computation of combinations. B. Sequential addition of analytes reveals their contribution to the ROC performance of the composite biomarkers. C. Stage-dependent performance of the composite biomarkers over stages I-IV. D. Boxplots of measured plasma concentrations for the analytes used in the composite biomarkers.

**Table 3.** Single analyte and composite plasma biomarkers for diagnosis of cancer compared to the ROC AUC of different benchmarks.

| Criterion<sup>a</sup> | CRC | LuCa | OvCa |
| --- | --- | --- | --- |
| Number of analytes | 10 | 22 | 13 |
| Superior to biomarker with |  |  |  |
| AUC 0.82 | 0 | 0 | 1 |
| AUC 0.88 | 0 | 0 | 1 |
| AUC 0.93 | 0 | 0 | 0 |
| Evaluated combinations of 2 | 90 | 462 | 156 |
| Superior to biomarker with |  |  |  |
| AUC 0.82 | 0 | 3 | 29 |
| AUC 0.88 | 0 | 0 | 24 |
| AUC 0.93 | 0 | 0 | 2 |
| Evaluated combinations of 3 | 240 | 3080 | 572 |
| Superior to biomarker with |  |  |  |
| AUC 0.82 | 0 | 60 | 156 |
| AUC 0.88 | 0 | 0 | 132 |
| AUC 0.93 | 0 | 0 | 19 |
| Evaluated combinations of 4 | 420 | 14630 | 1430 |
| Superior to biomarker with |  |  |  |
| AUC 0.82 | 2 | 783 | 523 |
| AUC 0.88 | 0 | 0 | 440 |
| AUC 0.93 | 0 | 0 | 37 |

<sup>a</sup>The optimal ROC AUC was computed for all combinations of 2, 3 or 4 significantly altered analytes per tumor type (10 for CRC, 22 for LuCa, 13 for OvCa). For a combination of $k$ analytes we computed $2^{k+1}$ classification scenarios: each analyte could be up- or downregulated and the analytes combined using logical AND or logical OR. For each classification scenario exhaustive search is used to check all possible cutoff values and optimize the ROC curve, i.e. finding highest possible true positive rate for every false positive rate yielding the highest possible AUC. Each combination was checked for superiority at the 99% significance level (Bonferroni correction) to biomarkers on par with the predicate devices Epi proColon (AUC 0.82), FIT (AUC 0.88) and ColoGuard (AUC 0.93).

### Tumor-stage-dependent biomarkers

In addition to early cancer detection, we also wanted to identify biomarkers that can discriminate cancer stage. We therefore evaluated correlations between protein and metabolite concentrations and stage for CRC, LuCa and OvCa respectively. We analyzed the 818 cancer cases and 119 non-cancer controls by Spearman rank correlation test, including all measured proteins and metabolites since the pre-analytical effects from the healthy control cohort comparisons are not an issue here. We found 4 (2 proteins, 2 metabolites) analytes for CRC, 111 (27 proteins, 84 metabolites) analytes for LuCa and 23 (12 proteins, 11 metabolites) analytes for OvCa that significantly correlated with stage (95% significance level, Bonferroni correction), with the MDK protein, and the two metabolites XS-VLDL-PL % and L-LDL-C % overlapping for all the three tumor types (Figure 4A-B, Supplementary Table 18). Next, we searched for single analytes that could discriminate stages III-IV from stages I-II tumors with statistical significance by computing ROC AUC, and found 0 analytes for CRC, 110 analytes (27 proteins, 83 metabolites) for LuCa and 8 analytes (4 proteins, 4 metabolites) for OvCa (2-sided Mann-Whitney *U* test under Bonferroni correction; Figure 4C-D; Supplementary Table 19). In the same way, we also searched for single analytes that could distinguish stage IV from stages I-III and found 22 (1 protein, 21 metabolites) for CRC, 78 (17 proteins, 61 metabolites) for LuCa, and 8 (4 proteins, 4 metabolites) for OvCa. From the single analytes that could separate stage IV cancers from lower stages, we computed all possible 4-combination composite biomarkers and the top combinations included upregulated CEACAM5, XL-VLDL-PL %, GlycA and downregulated L-LDL-CE % for CRC (ROC AUC 0.87), upregulated MUC16, DKK, KRT19 and downregulated Albumin for LuCa (ROC AUC 0.92), and upregulated MUC16, XXL-VLDL-PL %, CHI3L1 and downregulated Albumin for OvCa (ROC AUC 0.88) (Figure 4E, Supplementary Tables 20-22). Downregulated albumin in LuCa and OvCa is consistent with the hypoalbuminemia commonly observed in wide-spread carcinosis. Notably, the discrimination of stage IV CRC from stage I-III CRC was much better compared to the discrimination of stages III-IV from stage I-II. This is in contrast to LuCa and OvCa, where there was similar performance for stage IV vs I-III and stage III-IV vs I-II (Figure 4D). This is however consistent with our earlier observation that no single analytes were statistically significant for separation of CRC stage III+IV from stage I+II. Taken together, blood-based resolution of tumor stage based on the proteins and metabolites measured was more effective for LuCa and OvCa than for CRC, and metabolites contributed to the performance of several of the best composite biomarkers.

**Figure 4.**
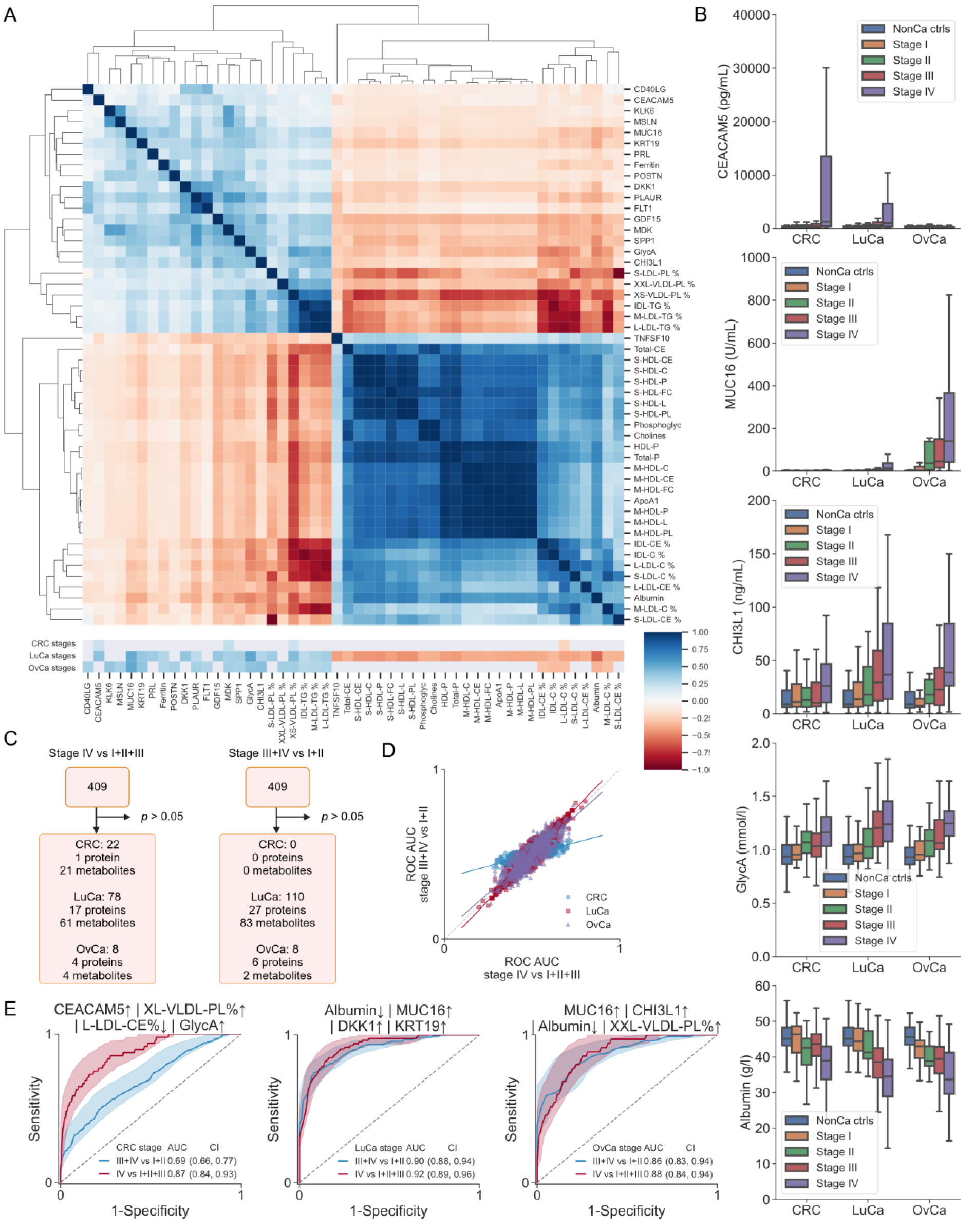
Tumor-stage-dependent proteomic and metabolomic signatures. A. Clustered heatmap of Spearman rank correlations between analytes and stages for CRC, LuCa and OvCa, respectively. Lower heatmap displays for which type of cancer the analytes had significant correlation with stage. Significant correlations with stage were observed for 4 proteins in CRC, 111 in LuCa and 23 in OvCa (adjusted *p* < 0.05). B. Boxplots of analyte concentrations for different tumor types and stages. C. Identification of analytes of all 409 (165 proteins and 244 metabolites) measured that could separate stage IV from stages I-III using the Mann-Whitney *U* test as well as analytes that could separate stages III-IV from stages I-II. D. ROC AUC values for separating stage IV from stages I-III, plotted against ROC AUC values for separating stage III-IV from stages I-II for all 409 measured single analytes. E. The ROC for the top combinations of composite biomarkers separating stages III-IV from stages I-II as well as separating stage IV from stages I-III. For each combination of selected analytes evaluated, we optimized the ROC curve by exhaustive search on all possible cutoff values combined either by logical OR (|) or logical AND (&).

### External validation of proteomic biomarkers

Finally, we performed external validation of individual proteins and selected composite biomarkers for detection of cancer vs healthy controls (Figure 5). To this end, we compared the data from stage I-III cancers and healthy controls in this study to data from the CancerSeek study^10^ where Luminex technology was used to measure one-quarter of the proteins analyzed here (Figure 5A). We found several single-protein biomarkers, including MUC-16/CA125 and GDF15, that had highly similar ROC AUC values between our study and the CancerSeek study, whereas some were slightly superior in CancerSeek (SPP1/OPN) or in this study (PECAM-1), respectively (Figure 5B, Supplementary Table 23). In all, 32 proteins were measured in both studies and after filtering analytes with technical issues we selected proteins with consistent behavior for each diagnosis. When 4-protein composite biomarkers were generated using the 8 retained proteins for CRC, 7 retained proteins for LuCa and 13 retained proteins for OvCa -, similar ROC performance was achieved in both datasets (Figure 5C, Supplementary Tables 24-26). In conclusion, we observed a high degree of similarity for biomarkers that we sought to validate in an independent data set, indicating that composite protein biomarkers can form the bases of robust and reproducible diagnostic tests for cancer detection.

**Figure 5.**
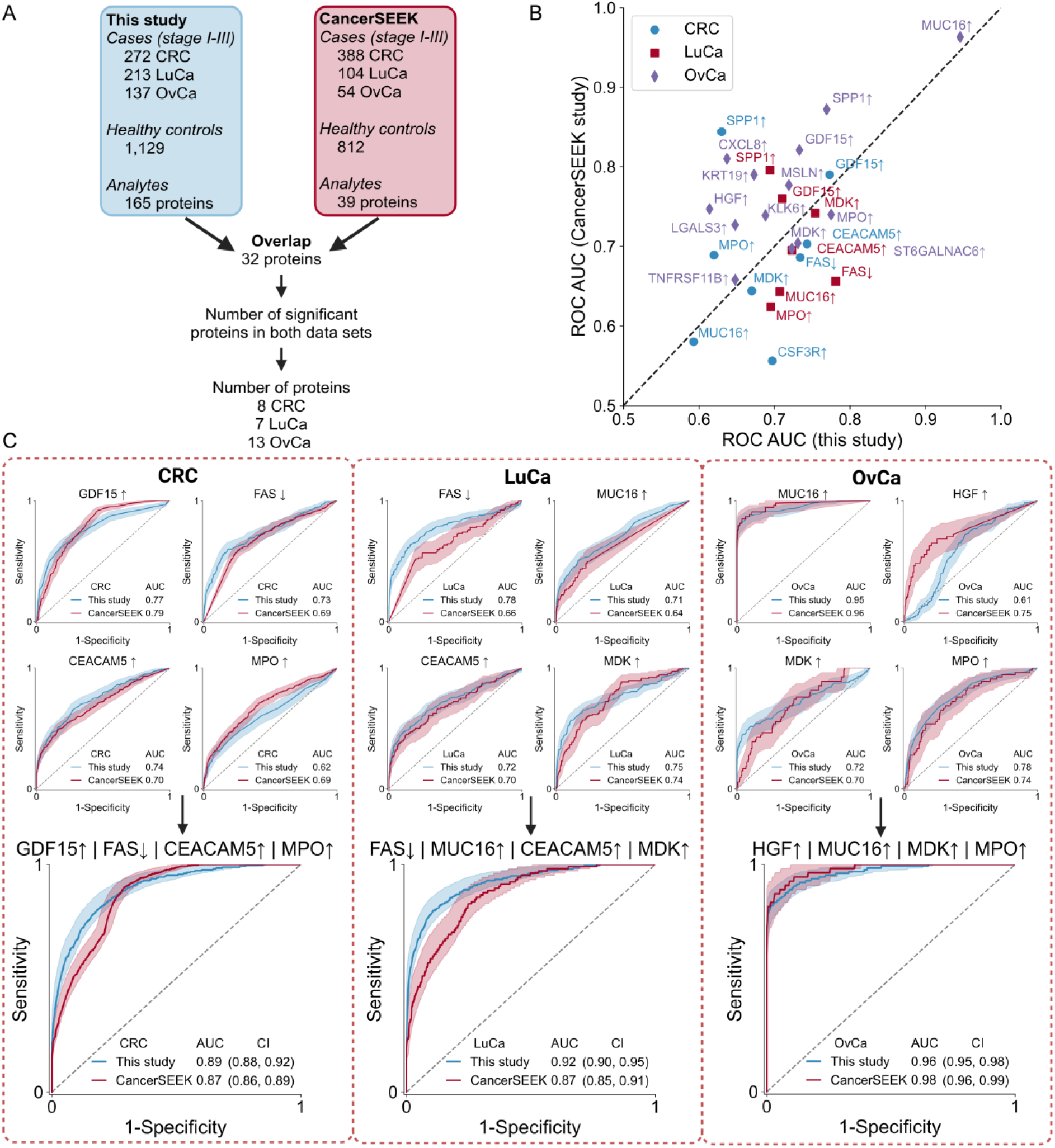
Validation of proteomic biomarkers for cancer detection in an external dataset. For a subset of proteins, external validation could be performed in CancerSEEK data. A. Cohort sizes and overlap in measured proteins in this study and in the CancerSEEK study. In total, 32 proteins were measured in both studies and 3 of these were excluded due to technical issues. Next, we enriched for proteins with consistent behavior, selecting for each tumor type those with ROC AUC > 0.5 for cancer stage I-III vs healthy controls in both studies. This resulted in 8 CRC, 7 LuCa and 13 OvCa proteins considered informative in both studies. B. The ROC AUC in this study (discovery) plotted against the ROC AUC in CancerSEEK study (validation) for all proteins regarded informative after filtering. C. The best 4-combination biomarkers of informative proteins found in this study for CRC, LuCa and OvCa, respectively, and the corresponding performance in CancerSEEK. For each biomarker, the ROC curves and AUC of the individual proteins and the composite biomarker are shown.

## Discussion

Here, we analyzed a large dataset and identified combinations of 2-4 analytes/proteins that performed better than an FDA approved early cancer detection blood test. Based on FDA pre-approval decisions on biomarkers since 1981, the Epi proColon ROC performance of 0.82 can be considered as a bar to surpass for a new blood-based biomarker test for early detection of cancer to have potential for regulatory approval and wider clinical adoption. We here succeeded in identifying composite plasma all-protein biomarkers that exceed the selected benchmark for all three tumor types. These combinations merit further development and evaluation in cohorts collected in a healthcare triage setting as well as in a screening setting. In both CRC and LuCa we found biomarkers that were significantly superior to the performance of detection of CRC by Epi proColon. In OvCa, we identified biomarkers with ROC performance superior to the at the time of analyses most sensitive FDA approved early detection test for cancer currently available, which is the feces test Cologuard with ROC AUC 0.93 for detection of CRC. For OvCa, the OVA-1 test is an FDA-approved blood serum test for assessing risk for ovarian malignancy in women that have been previously diagnosed with an ovarian mass for which they are scheduled for surgery. However, OVA1 is not a test for early cancer detection in women that do not have an already detected ovarian mass, and is therefore not a suitable benchmark for this study. During 2024, past the analyses of this study, two new early detection tests for CRC obtained FDA premarket approval, the cfDNA blood-based Shield test, and the multitarget stool DNA test Cologuard Plus. Both these tests show higher sensitivity than the corresponding benchmark tests that we used here. In clinical evaluation, the Shield test had 83.1% sensitivity^52^, and Cologuard Plus 93.9% sensitivity^53^ for detection of CRC. Although the blood-based Shield test outperforms the previous state-of-art, the Epi proColon test, detection of early stage cancer and precancerous lesions remains a challenge, with sensitivity of the Shield test for clinical stage I cancer being 55% and for advanced precancerous lesions 13%^52^.

To the best of our knowledge, this is the first study integrating affinity proteomics and metabolomics to develop composite biomarkers. The cores of the composite cancer detection biomarkers identified here contained well-established single protein biomarkers such as MUC16/CA125 or CEACAM5/CEA, effectively supporting their relevance. In CRC, while FLT1 has previously been found elevated^54^, IL-19 has not. In LuCa, the PLAUR/suPAR is previously reported elevated in non-small cell LuCa (NSCLC) but not in small cell LuCa (SCLC)^55^, whereas elevated FNDC5/irisin has not previously been linked to LuCa. Consistent with our data, Midkine (MDK) has previously been found elevated in LuCa^56^ and OvCa^57^. An interesting observation is that, as compared to the literature, MUC16/CA125 performed extremely well for detection of OvCa in our data, and also in the external Luminex-based validation dataset from the CancerSeek study. While CA125 is in use in OvCa monitoring and prognostics, including as part of the OVA1 test^58, 59^, it has some issues that limit its clinical value. One example is that it was previously found not to be elevated in ∼20% of women with epithelial OvCa^60^. One potential explanation is that the antibodies to CA125 used in the Luminex assays perform better than those used in other affinity proteomics assays, which could potentially be utilized to improve current CA125-containing biomarkers. Interestingly, plasminogen (PLG) elevation, which has not previously been reported in OvCa, was part of strong biomarkers in combinations both with and without CA125. In contrast, both of the two other elevated plasma proteins in the best OvCa biomarker without Ca125, KLK6 and CCL2, have prior association with OvCa^61, 62^. For NMR metabolomics, this study encompasses twice the number of CRC cases^63, 64^, twice the number of LuCa cases^65^ and twice the number of OvCa cases^66^ than prior art. Several NMR analytes of interest here, including albumin and glycoprotein acetyls (GlycA), have regulatory approval for clinical use, which could enable implementation of composite biomarkers incorporating metabolites. However, the cohort differences impacted variability for metabolites more than was the case for proteins in this study, potentially because of differences in fasting status before blood draw^67^ between the two cohorts. While we did find metabolites potentially useful for stage determination here when we analyzed only the U-CAN cohort, the observation that only two metabolites passed filtering for pre-analytical differences between the two cohorts when U-CAN cases were compared to the healthy controls of the EpiHealth cohort suggests that NMR-based tests will require highly standardized sampling and handling for robustness of clinical implementations. Taken together, the best performing composite biomarkers for each tumor type combine known and novel biomarkers, supporting their relevance.

Another potential use of cancer biomarkers is to separate early from late stage tumors. Interestingly, a larger number of metabolites compared to proteins had statistically significant differences between late stages and early stages among the cancer cases. Some analytes, such as Albumin and GlycA, are well-known general disease markers and are likely not tumor specific. However, in combination with more tumor-specific proteins this could still be useful in a composite biomarker. For LuCa and OvCa, many analytes showed consistent increase or decrease with increasing stage (correlation analysis and box plots in Figure 4), whereas for CRC most analytes displayed a large difference between stage IV and stages I-III. For LuCa and OvCa, composite biomarkers for stage separation could discriminate stages III-IV from stages I-II with similar ROC AUC as discriminating stage IV from stages I-III, which was not the case for CRC, suggesting that stage IV CRCs but not stages I-III show strong circulating proteomic and metabolomic signatures (Figure 3C). This is consistent with the generally lower performance for CRC biomarkers compared to LuCa and OvCa discovered in this study. Overall, metabolites proved more useful for tumor stage discrimination than early detection.

The FDA approval process for a new biomarker is centered on comparing its sensitivity and specificity to that of biomarkers already approved by the regulator for the same purpose. Here, we used large numbers of samples and a novel computational approach^51^ to evaluate combinations of analytes, for each combination identifying the optimal direction of change for each analyte that produced the best ROC AUC performance. While partitioning of data for training, test and validation is current convention, we reasoned that external validation by analysis of independently generated data is more meaningful given that study-specific biases caused by pre-analytical sample handling and cohort composition are not corrected for by partitioning. After removing analytes with inflated ROC AUC likely due to pre-analytical differences between study cohorts, we used hypothesis testing under Bonferroni correction to determine if the combinations superseded clinical benchmarks tests in diagnostic performance. While a potential drawback of the filtering approach applied is the loss of biomarkers that also recognize non-malignant pre-cancerous lesions, we reasoned that it would be more useful to unambiguously identify biomarkers of malignant disease. While the approach used here identifies the combinations of analytes giving the best ROC performance, the optimal cut-off values need to be determined through analysis of additional data. To provide true external validation, we will next analyze the identified composite biomarkers in samples collected before diagnosis at suspicion of cancer as well as in a population screening setting, which represents the main use cases for diagnostic blood biomarkers. A potential use of biomarkers with high tumor-type specificity, as the ones identified here, is to be able to discriminate abdominal tumor types when patients present with diffuse symptoms that are not disease-specific. For example, patients with disseminated OvCa often present with similar symptoms and radiological findings as CRC patients. The discrimination of OvCa from CRC exemplifies such clinical needs, where a reliable blood test could be a supplement or even alternative for biopsy if validated clinically.

Since infrastructure and workflows are straightforward, cost effective and well established in clinical laboratories, protein-based diagnostic biomarkers for cancer may be preferred as compared to analyses of circulating DNA or RNA. However, the requirements for sensitivity and tumor type specificity are rarely fulfilled by single-analyte biomarkers, but demands a combination of several proteins, metabolites or nucleic acid targets^68^. For reliability and cost reasons, a diagnostic test based on such combinations should incorporate as few analytes as possible. We therefore capped combination sizes at 4 analytes in this study. Although this is a low number considering other prominent proteomics-containing cancer signatures^10^, we identified hundreds of potential tumor-type-specific biomarkers that perform at least equally well as the prior art. While there are other affinity proteomics technologies that enable measurement of thousands of proteins, and can thus provide much larger datasets for de novo biomarker discovery, the one used here is implemented on an FDA approved instrument platform, which can greatly facilitate clinical translation. Analysis of differences in diagnostic performance within and between study cohorts allowed selection of a limited set of biomarkers. In this study these biomarkers could either be shown to behave similarly versus non-malignant controls from both study cohorts, or be validated in an external cohort. However, as we measured 4-fold more proteins than the external dataset from the CancerSeek study, only a subset of combinations could be externally validated at this point. In comparison to CancerSEEK, several of the proteins we could compare had remarkably similar ROC AUC for cancer versus population controls, while others were slightly different. These differences can indicate sensitivity to pre-analytical variation, but could also be caused by e.g. the different age distributions of cases and controls in CancerSEEK.

## Methods

### Ethical approval

The study was approved by the Swedish Ethical Review Authority (EPM dnr 2019-00222) with amendments. The research was in line with informed consents obtained from patients included in U-CAN^24^ (EPN Uppsala 2010-198 with amendments) and research participants in EpiHealth^25^.

### Cases and controls

Study size was determined according to (Ekström et al, manuscript in preparation). Cancer cases were selected from the full population of individuals included 2010-2020 in the longitudinal prospective cohort U-CAN^24^ at Uppsala University Hospital. Inclusion criteria for this study for cancer cases included sample freezing time ≤4 h after blood draw, registration in the national quality registry for the respective cancer diagnosis (the Swedish Colorectal Cancer Registry, the Swedish National Quality Registry for Lung Cancer, and the Swedish Quality register for Gynecologic Cancer), age at diagnosis range 45-75 (CRC and LuCa) or 40-80 (OvCa), no non-surgical or surgical cancer treatment before blood sampling, sampling within 90 d of the diagnosis date stated in the respective national quality registry, no known concurrent or prior malignancy reported in the Swedish Cancer Registry, and availability of sufficient plasma samples. Epithelial borderline ovarian tumors (n=24) were included among the OvCa cases. As technical controls for pre-analytical handling, we used plasma samples from patients in U-CAN with non-malignant or pre-malignant conditions in the respective organ sampled under identical handling conditions as the cancer cases. The most common benign conditions were colon polyps, adenomas, prolapse and incontinence; gynecological cyctoadenomas of various histology, endometriosis, endometrial hyperplasia and polyps, myomas and cervical intraepithelial neoplasia (CIN); chronical inflammatory conditions of the lung. The EDTA plasma samples from U-CAN participants had been drawn non-fasting at diagnosis before administration of any anti-cancer treatment, and subjected to central standardized handling at RT followed by freezing for biobanking at -80 °C. Population controls were selected from individuals included in the prospective study EpiHealth^25^ in 2010-2015, that had not been registered in the Swedish Cancer Registry as having any type of cancer up to 5 years after sampling. The EpiHealth individuals had been sampled for blood at a test center at Uppsala University following 6 h fasting, with plasma centrifuged and stored for up to 6 h at +4 °C followed by freezing for biobanking at -80 °C.

### Measurements of proteins and metabolites

Plasma levels of 165 different proteins (Supplementary Table 27) were measured using a suspension array ELISA system (BioPlex 200, Bio-Rad) and 10 antibody panels (Human Circulating Cancer Biomarker Panel 1, Human Circulating Cancer Biomarker Panel 3, Human Circulating Cancer Biomarker Panel 4, Human Cancer metastasis Biomarkers panel, Human Immuno-Oncology Checkpoint Protein Panel 1, Angiogenesis Panel 2, Human Cytokine/Chemokine Panel, Human Cytokine/Chemokine Panel II, Human Cytokine/Chemokine Panel IV, Human Myokine Panel) according to the manufacturer’s instructions. Using xPONENT® software (ThermoFisher), data was collected from 50 beads per bead set for all 10 kits. The OvCa marker WFDC2 (HE4) was measured but excluded for technical reasons (assay failure) after consulting the manufacturer. The plasma levels of 244 metabolite parameters (Supplementary Table 28) were determined by NMR using the Blood analysis service by Nightingale Health, Helsinki, Finland.

### Data analysis

For all analytes we computed ROC AUC for distinguishing cancer from non-cancer controls and cancer from healthy controls. Based on extensive study design simulations, all data was used for validation. All data points were included in ROC computations and statistical analyses. In boxplot visualizations, the whiskers extend to 1.5 times the interquartile range, and outliers are not shown due to distortion of scales. Statistical significance at the 95% level was tested using two-sided Mann-Whitney U test and Bonferroni correction for multiple hypothesis testing. The number of hypotheses tested and corrected for was 409*3 = 1227. Analytes that were significant both versus non-cancer controls and healthy controls were included in computation of combination biomarkers. Data analysis and visualization were done in Python and corresponding data science packages (*Python 3.9.18, numpy 1.21.4, pandas 1.3.4, matplotlib 3.5.0, seaborn 0.13.0, plotly 5.6.0, scikit-learn 1.2.1, networkx 3.1*). Plots were generated using Python and figure panels composed with Inkscape and biorender.com.

### Composite biomarkers for cancer detection

We used a novel algorithm to compute the ROC curves for combinations of up to 4 analytes while optimizing the cut-off values for each analyte^51^. For a combination of *k* analytes we computed *2^k+1^* classification scenarios: each analyte can be up- or downregulated and combined using logical AND or logical OR. For each classification scenario exhaustive search was used to check all possible cutoff values and optimize the ROC curve, i.e. finding highest possible true positive rate for every false positive rate yielding the highest possible AUC. We designed the study according to results from prior simulations in this statistical framework that revealed that the most efficient use of cases and controls given the number of hypotheses tested was to assign all to a validation step (J. Ekström et al, Manuscript in preparation). We computed the ROC AUC for separating cancer cases of stages I-IV from the healthy controls of the population cohort given the over/under rules identified in the first step but allowing for optimization of new cut-off values. We then performed statistical hypothesis testing of whether the biomarker separated cases from controls better than a benchmark biomarker with the same properties as the currently best FDA approved diagnostic blood test for cancer, Epi proColon (ROC AUC 0.82), and the two best feces-based tests Fecal Immunochemical Test FIT (ROC AUC 0.88) or Cologuard (ROC AUC 0.93) at the 99% significance level given the same over/under rules that were generated in the comparison of cancers to non-malignant disease controls. This process was repeated for each tumor type. We used Bonferroni correction to adjust p-values and corrected for all computed combinations of respective order. Thus, we evaluated in total 708 2-combinations, 3,892 3-combinations and 16,480 4-combinations.

### Biomarkers for stage separation

The correlation analysis was performed using two-sided Spearman rank correlation test and Bonferroni correction for multiple hypothesis testing (409 hypotheses tested) to find analytes that were statistically significantly correlated with stage. Two-sided Mann-Whitney U test and Bonferroni correction was used to find individual analytes that could discriminate later stages from earlier stages: stage IV vs stages I-III and stages III-IV vs stages I-II. From the significant analytes we computed composite biomarkers in the same way as described above.

### External validation of protein biomarkers

For external validation of protein biomarkers, we computed ROC AUC for combinations using the same rule set as above and data from the CancerSeek study, which contains 388 CRC, 104 lung, and 54 OvCa of stages I-III along with 812 healthy controls sampled in the same pre-analytical handling^10^. For the proteins measured in both studies we used the same methodology and performed the same analysis as described above. First, we identified analytes that were statistically significant, computed all possible 4-combination composite biomarkers and compared performance in the two datasets. When computing the composite biomarkers, the cutoffs were optimized in the two datasets separately. This means that the same behavior (up/down-regulation) and combination of proteins were compared but not absolute cutoff values. This is to circumvent baseline differences due to differences in experimental protocol and sample handling.

## Data availability

The datasets generated in the study for cases and non-cancer controls from the U-CAN cohort are available in the SciLifeLab Data Repository, with DOI: 10.17044/scilifelab.28351367. Datasets generated for EpiHealth healthy controls are not publicly available due to limitations with the informed consent, however, are available from the corresponding author on reasonable request.

## Code availability

All code used to generate the figures can be obtained with a reasonable request to the corresponding author.

## Data Availability

All data produced in the present study are available upon reasonable request to the authors.

https://www.science.org/doi/suppl/10.1126/science.aar3247/suppl_file/aar3247_cohen_sm_tables-s1-s11.xlsx

## Acknowledgments

The study was supported by grants to T.S. from the Swedish Cancer Society (CAN 2018/772, 21 1719 Pj, 2024 3831 Pj), the Sjöberg Foundation and Lena Wäppling’s foundation, and from the Swedish Cancer Society to J.Å.Ö. (CAN 21-0430-PT) and B.G. (190382PJ01H). We thank U-CAN for access to blood samples collected through Uppsala Biobank, Uppsala University Hospital and Uppsala University with support from the Swedish Government (SRA grant CancerUU), and EpiHealth for blood samples collected with support from the Swedish Government through the Swedish Research Council.

