## Supplemental_material for "Composite proteomic and metabolomic plasma biomarkers for detection of colorectal, lung and ovarian cancers"

### Supplementary Materials

**Supplementary tables**

Table S1. ROC AUC and p-values for all measured unitary proteomic and metabolomic biomarkers in different cancer types, pan-cancer, and non-cancer controls versus healthy controls.

Table S2. All significant (p<0.05) unitary biomarkers for separating CRC from non-cancer controls.

Table S3. All significant (p<0.05) unitary biomarkers for separating LuCa from non-cancer controls.

Table S4. All significant (p<0.05) unitary biomarkers for separating OvCa from non-cancer controls.

Table S5. All significant (p<0.05) unitary biomarkers for separating Pan-cancer from non-cancer controls.

Table S6. All significant unitary biomarkers for separating CRC from non-cancer controls (p<0.05) as well as from healthy controls (p<0.05).

Table S7. All significant unitary biomarkers for separating LuCa from non-cancer controls (p<0.05) as well as from healthy controls (p<0.05).

Table S8. All significant unitary biomarkers for separating OvCa from non-cancer controls (p<0.05) as well as from healthy controls (p<0.05).

Table S9. All significant unitary biomarkers for separating Pan-cancer from non-cancer controls (p<0.05) as well as from healthy controls (p<0.05).

Table S10. Composite 4-analyte biomarkers for CRC vs healthy controls significantly (p<0.01) better than a biomarker with ROC AUC 0.82.

Table S11. Composite 2-analyte biomarkers for LuCa vs healthy controls significantly (p<0.01) better than a biomarker with ROC AUC 0.82.

Table S12. Composite 3-analyte biomarkers for LuCa vs healthy controls significantly (p<0.01) better than a biomarker with ROC AUC 0.82.

Table S13. Composite 4-analyte biomarkers for LuCa vs healthy controls significantly (p<0.01) better than a biomarker with ROC AUC 0.82.

Table S14. Unitary biomarker for OvCa vs healthy controls significantly (p<0.01) better than a biomarker with ROC AUC 0.88.

Table S15. Composite 2-analyte biomarkers for OvCa vs healthy controls significantly (p<0.01) better than biomarkers with ROC AUC 0.82, 0.88 and 0.93.

Table S16. Composite 3-analyte biomarkers for OvCa vs healthy controls significantly (p<0.01) better than biomarkers with ROC AUC 0.82, 0.88 and 0.93.

Table S17. Composite 4-analyte biomarkers for OvCa vs healthy controls significantly (p<0.01) better than biomarkers with ROC AUC 0.82, 0.88 and 0.93.

Table S18. Spearman rank correlations with stages 0-4, all unitary analytes for CRC, LuCa and OvCa respectively. Significant analytes (p<0.05) are highlighted green.

Table S19. ROC AUC and p-values for all measured unitary proteomic and metabolomic biomarkers for separating stages in CRC, LuCa, OvCa and pan-cancer.

Table S20. Composite 4-analyte biomarkers for separating CRC stages.

Table S21. Composite 4-analyte biomarkers for separating LuCa stages.

Table S22. Composite 4-analyte biomarkers for separating OvCa stages.

Table S23. ROC AUC and p-values for all unitary analytes measured both in the CancerSeek study and in this study.

Table S24. Composite 4-analyte biomarkers for CRC vs Healthy ctrls in this study and CancerSeek.

Table S25. Composite 4-analyte biomarkers for LuCa vs Healthy ctrls in this study and CancerSeek.

Table S26. Composite 4-analyte biomarkers for OvCa vs Healthy ctrls in this study and CancerSeek.

**Supplemental figures**

Supplementary Figure 1. Population controls and cancer cases.

Supplementary Figure 2. Pre-malignant and non-malignant disease control samples.

Supplementary Figure 3. Characteristics of the colorectal cancer patients in the study.

Supplementary Figure 4. Characteristics of the lung cancer patients in the study.

Supplementary Figure 5. Characteristics of the ovarian cancer patients in the study.

#### Supplementary Figure 1

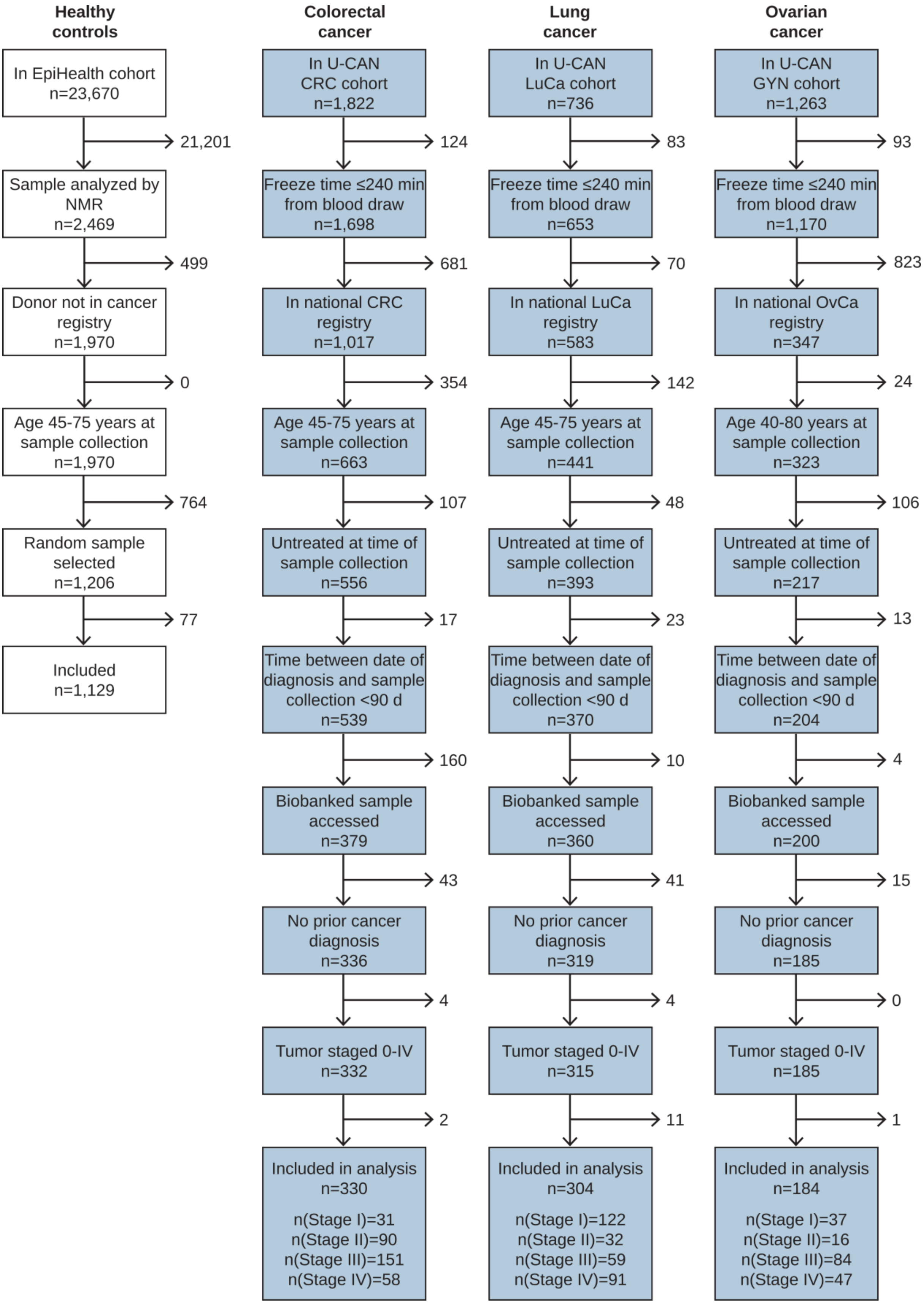

**Supplementary Figure 1. Selection of population controls and cancer cases.** Population controls were obtained from the EpiHealth study, which recruited 23,670 healthy individuals aged 45-75 living in Uppsala County, Sweden, during 2010-2015 (23435790). A subset of 2,469 individuals had already been characterized by other -omics methods. From these, 1,970 had not had a malignant disease at least 5 years after inclusion in EpiHealth, as determined by not being recorded in the national Swedish Cancer Registry. A random sample of 1,206 individuals was drawn and 1,129 were analyzed with Luminex proteomics and included as a healthy population control group. Cancer patients diagnosed during 2010-2020 in Uppsala County were obtained from the prospective longitudinal cohort U-CAN. Inclusion criteria for this study included complete reporting to the respective Swedish national quality registry, blood sample frozen within 4 h of blood draw, age 45-75 (CRC and LuCa) or 40-80 (OvCa) years at sample collection, no surgical or non-surgical cancer treatment before sampling, sample drawn within 90 days of diagnosis, and no other concurrent or prior cancer. Patients with cancer stage I-IV were selected as cases.

**Supplementary Figure 2**

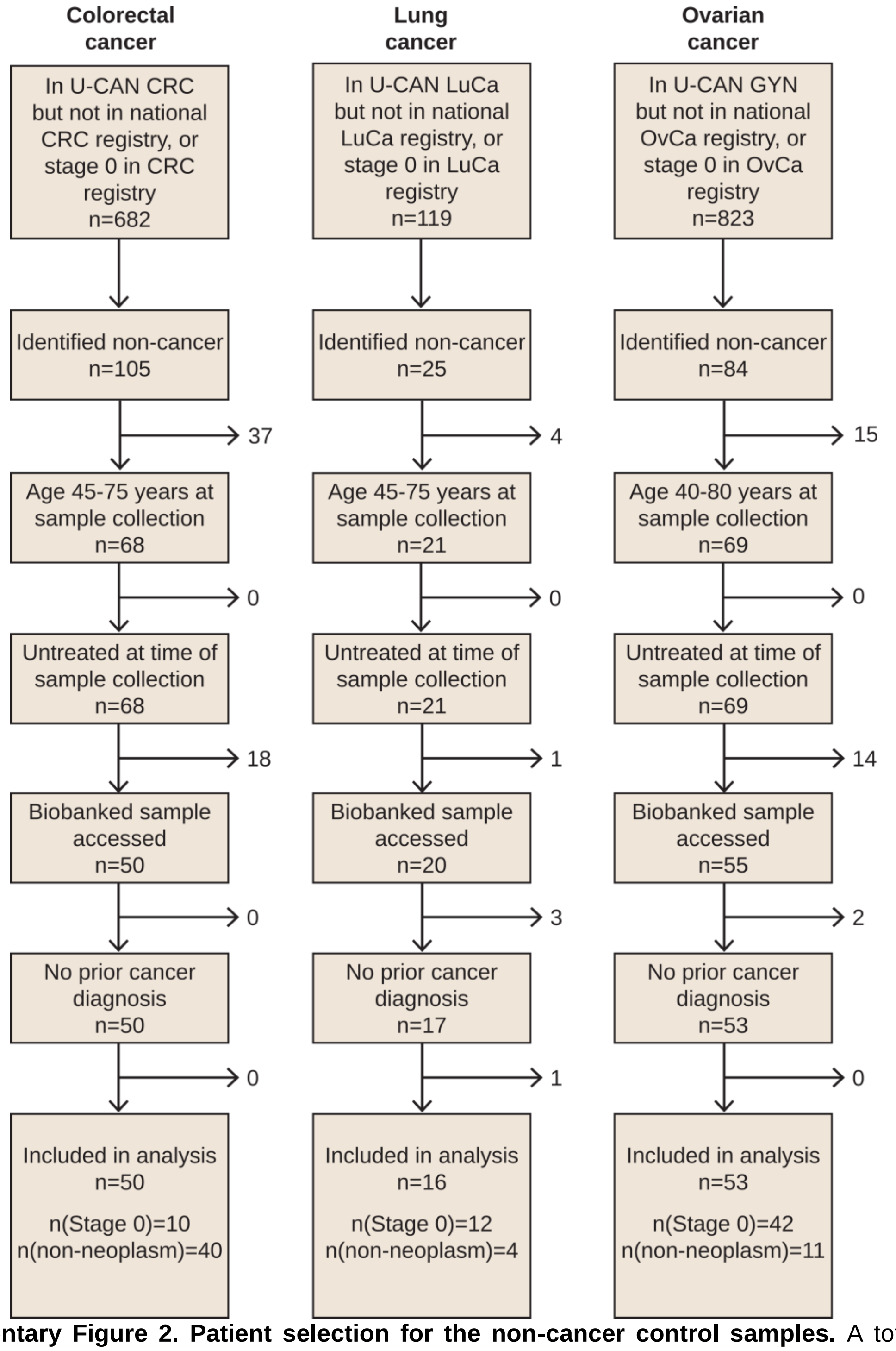


**Supplementary Figure 2. Patient selection for the non-cancer control samples.** A total of 119 controls, of which 64 were stage 0 pre-malignant tumors and 55 were from patients with non-malignant disease, were selected from the U-CAN cohort. These control patients had been included in U-CAN on suspicion of colorectal, lung or gynecological cancer, respectively, but were eventually diagnosed with a non-malignant or pre-malignant (stage 0) condition. The blood samples were collected with identical pre-analytical handling as for the cancer cases. The inclusion criteria for this study included the blood sample being frozen within 4 h of blood draw, age 45-75 (CRC and LuCa) or 40-80 (OvCa) y at sample collection, no surgical or non-surgical cancer treatment before sampling, and no concurrent or prior cancer diagnosis. The vast majority of controls (114 of 119) were not included in the Swedish national quality registries for CRC, LuCa or OvCa, however five stage 0 patients were.

#### Supplementary Figure 3

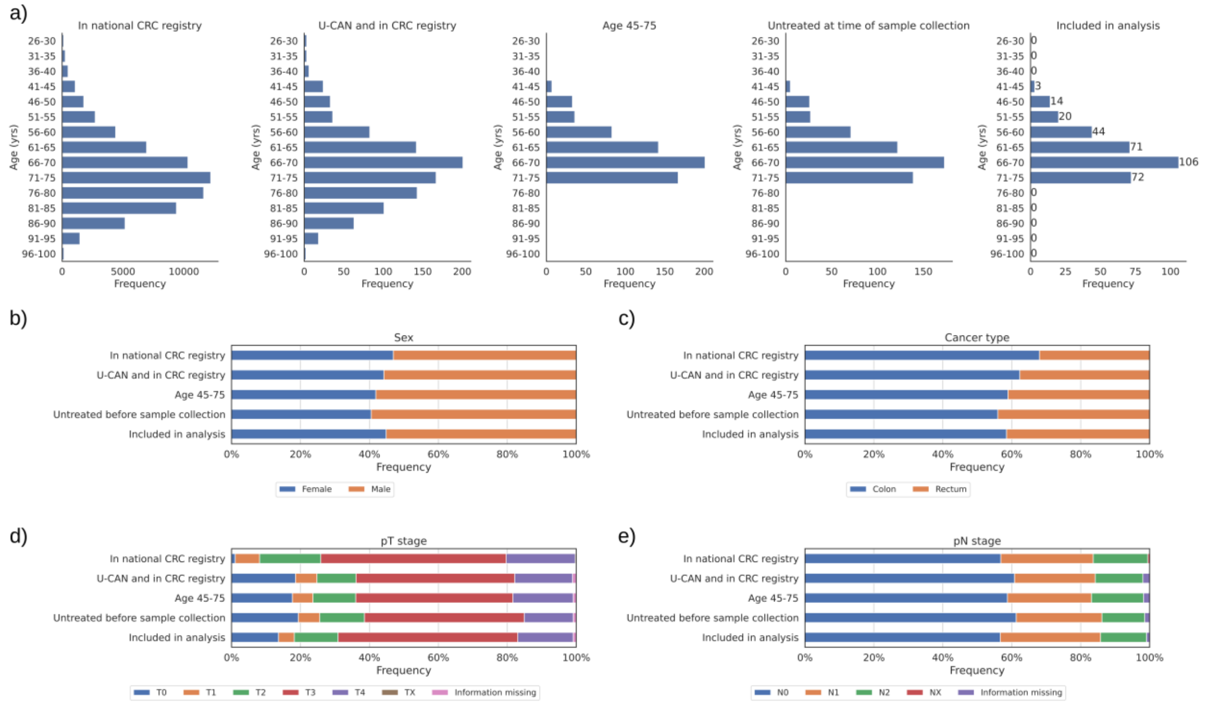


**Supplementary Figure 3**. **Characteristics of the colorectal cancer patients in the study.** Patient level data were obtained from the Swedish National quality registries for colon and rectal cancer for all patients included in the U-CAN CRC cohort. To enable comparison to the lung cancer patient population in Sweden, aggregated data for the whole population in the age range 26-100 years that was reported in the colon and rectal cancer quality registries during years 2010-2020 were obtained from the registries' public interactive reports (https://statistik.incanet.se/kolorektal/kolon/, and https://statistik.incanet.se/kolorektal/rektum/; accessed 12 Jan 2023). The distribution of cases with respect to (a) age, (b) sex, (c) cancer type, (d) pT stage, and (e) pN stage at diagnosis or surgery of the primary tumor is shown at key steps that were applied for selection of the final study cohort. In national CRC registry, n=68,068; U-CAN and in CRC registry, n=1,017; Age 45-75 years, n=663; Untreated before sample collection, n=556; Included in analysis, n=332. For pT and pN stage, data for information missing is not available in the public interactive reports, thus the group In national CRC registry is n=52,349 and n=52,288, respectively for pT and pN stage. For the patients were the quality registry gave T0 for pT stage, the cT stage and patient medical records were used for assessment of the tumor stage at diagnosis for the final study cohort since these patients were found to have responded to pretreatment that was given after the blood sample was donated, but before surgery. CRC, colorectal cancer; pT/pN, pathological T/N stage; TX, the primary tumour cannot be assessed; NX, regional lymph nodes cannot be assessed.

#### Supplementary Figure 4

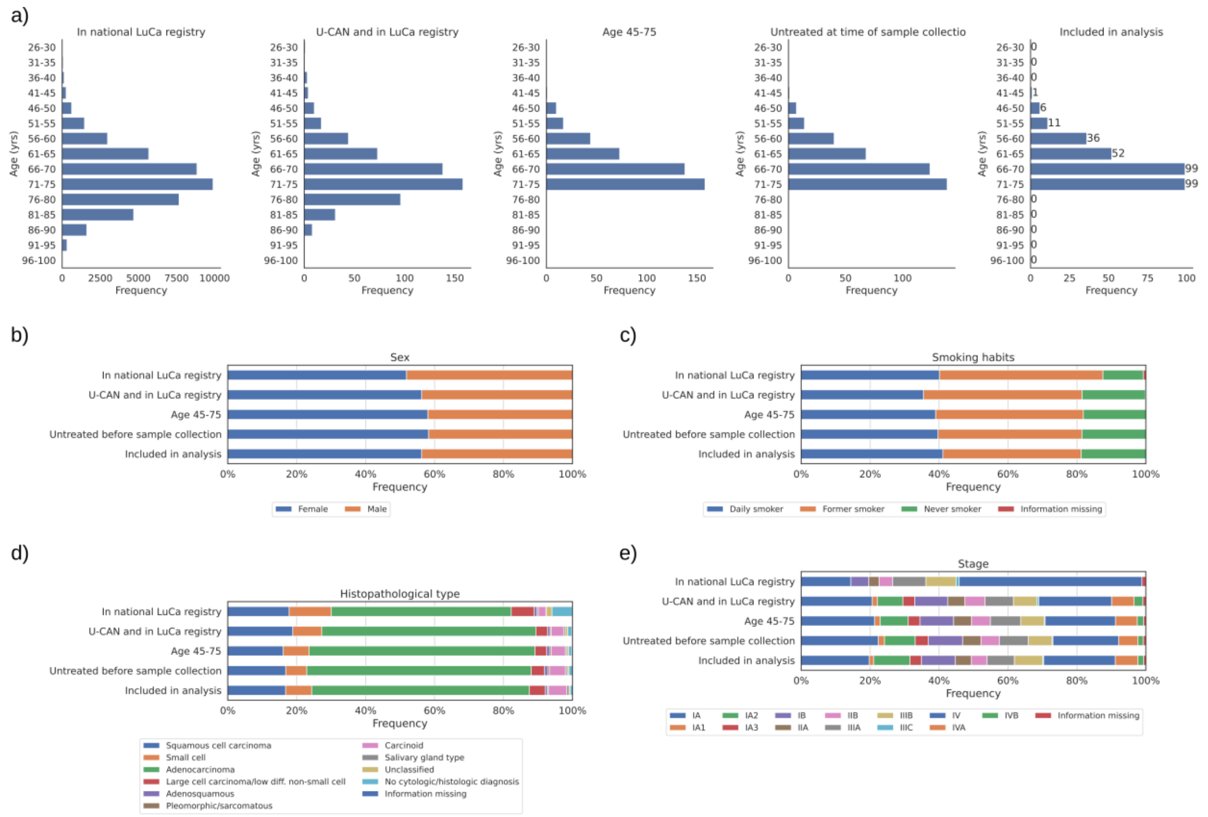


**Supplementary Figure 4**. **Characteristics of the lung cancer patients in the study.** Patient level data were obtained from the Swedish National quality registry for lung cancer for all patients included in the U-CAN lung cancer cohort. To enable comparison to the lung cancer patient population in Sweden, aggregated data for the whole population in the age range 26-100 years that was reported in the lung cancer quality registry during years 2010-2020 were obtained from the registry's public interactive report (https://statistik.incanet.se/Lunga/; accessed 12 Jan 2023). The distribution of cases with respect to (a) age, (b) sex, (c) smoking habits, (d) histopathological type of the primary tumour, and (e) disease stage at diagnosis or surgery of the primary tumor is shown at key steps that were applied for selection of the final study cohort. In national LuCa registry, n=44,306; U-CAN and in LuCa registry, n=583; Age 45-75 years, n=441; Untreated before sample collection, n=393; Included in analysis, n=315. LuCa, lung cancer.

#### Supplementary Figure 5

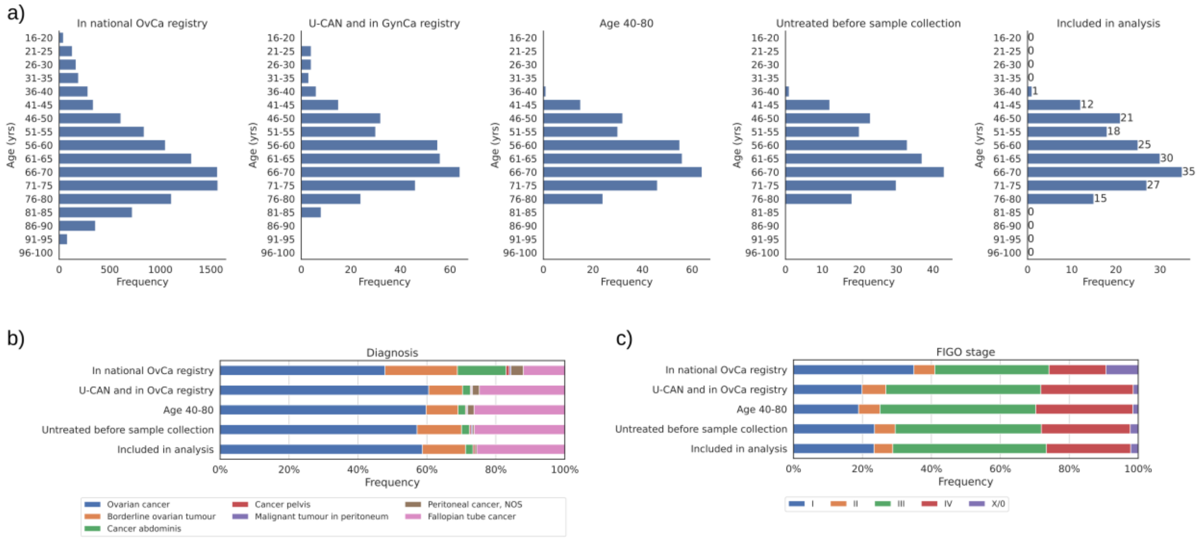


**Supplementary Figure 5**. **Characteristics of the ovarian cancer patients in the study.** Patient level data were obtained from the Swedish National quality registry for gynecological cancer for all patients included in the U-CAN gyn cancer cohort. To enable comparison to the ovarian cancer patient population in Sweden, aggregated data for the whole population in the age range 16-100 years that was reported in the gynecological cancer quality registry, sub-registry for ovarian cancer, during years 2012-2020 were obtained from the registry public interactive report (https://statistik.incanet.se/gyncancer/; accessed 12 Jan 2023). The distribution of cases with respect to (a) age, (b) diagnosis, and (c) FIGO stage at diagnosis or surgery of the primary tumor is shown at key steps that were applied for selection of the final study cohort. In national OvCa registry, n=10,374; U-CAN and in OvCa registry, n=347; Age 40-80 years, n=323; Untreated before sample collection, n=217; Included in analysis, n=185. OvCa, ovarian cancer; NOS, not otherwise specified.
